# Semaglutide for Weight Loss in Patients with a Family History of Thyroid or Uterine Cancer: A Systemic Review

**DOI:** 10.1101/2025.02.27.25323032

**Authors:** Mohammed Munshi, Sabrina Jahangir

**Affiliations:** Department of Public Health, Daffodil International University, Dhaka, Bangladesh; Department of Medicine, Shaheed Suhrawardy Medical College and Hospital, Dhaka, Bangladesh

**Author notes:** Corresponding Author: Name: Mohammed Munshi, Postal Address: House-6, Lane-21, Block-C, Avenue-5, Mirpur-11Pallabi, Dhaka-1216, Bangladesh, Contact No.

**Keywords:** Semaglutide, Thyroid Cancer, Uterine Cancer, Weight Loss, GLP-1 Receptor Agonists

## Abstract

**Background:** The weight loss medication semaglutide has recently become widely recognized for its effectiveness. Medical professionals have expressed safety concerns about semaglutide for patients who have a family background of thyroid or uterine cancers. The FDA has communicated about potential dangers requiring careful selection of patients before treatment. This study reviews existing research about semaglutide as a weight management treatment for patients with familial cancer histories while analyzing its potential advantages and dangers.

**Objective:** To examine recent studies on the use of semaglutide for weight loss in individuals with family histories of thyroid or uterine cancers, while also considering both potential risks and benefits.

**Primary & Secondary Outcome Measures:** Semalutide’s safety concerns are a concern for patients with family history of thyroid or uterine cancer. Semaglutide has potential medical advantages such as promoting weight loss and decreasing cancer risk, which is its secondary effect.

**Intervention:** Semaglutide is now a widely used treatment for weight loss, acting as an antagonist for the GLP-1 receptor. Primary & Secondary Outcome Measures: Semalutide’s safety concerns are a concern for patients with family history of thyroid or uterine cancer. Semaglutide has potential medical advantages such as promoting weight loss and decreasing cancer risk, which is its secondary effect.

**Methods:** A systematic literature search was conducted to identify studies investigating the use of semaglutide in patients with a family history of thyroid or uterine cancer. The following databases were searched: PubMed, Google Scholar, and ClinicalTrials.gov. Search terms included “semaglutide,” “thyroid cancer,” “endometrial cancer,” “GLP-1 receptor agonists,” and “weight loss.” Studies published between 2010 and 2024 were included. The search strategy was designed to capture both preclinical and clinical studies. Studies were included if they investigated the safety or efficacy of semaglutide in patients with a family history of thyroid or uterine cancer and provided clear data on cancer risk or weight loss outcomes. Studies lacking methodological rigor or not addressing the research question were excluded. The risk of bias in included studies was assessed using the Cochrane Risk of Bias Tool. Due to the heterogeneity of the studies, results were synthesized narratively, and a meta-analysis was not performed.

**Results:** A total of 4 studies involving 1,699,198 participants were included. Results showed no significant link between semaglutide use and thyroid cancer risk in human studies, although rodent studies indicated a higher risk of thyroid C-cell tumors. Preclinical data suggested potential benefits of semaglutide in reducing endometrial cancer risk, but definitive human research is needed.

**Article Summary:** *Strengths and limitations of this study:* This review utilizes a systematic and comprehensive search strategy across multiple databases, focusing on high-risk populations and applying clear inclusion criteria for both preclinical and clinical data. However, the studies included are heterogeneous, human data on cancer risk is limited, long-term safety information is lacking, potential publication bias may be present, and the findings cannot be generalized.

*Conclusion:* It is recommended that healthcare providers carefully review patient records and exercise caution when prescribing semaglutide to those with a history of thyroid or uterine cancers in their family.

## INTRODUCTION

The weight loss medication semaglutide has recently become widely recognized for its effectiveness. Medical professionals have expressed safety concerns about semaglutide for patients who have a family background of thyroid or uterine cancers. The FDA has communicated about potential dangers requiring careful selection of patients before treatment. This study reviews existing research about semaglutide as a weight management treatment for patients with familial cancer histories while analyzing its potential advantages and dangers.

## METHODS

A systematic literature search was conducted to identify studies investigating the use of semaglutide in patients with a family history of thyroid or uterine cancer. The following databases were searched: PubMed, Google Scholar, and ClinicalTrials.gov. Search terms included “semaglutide,” “thyroid cancer,” “endometrial cancer,” “GLP-1 receptor agonists,” and “weight loss.” Studies published between 2010 and 2024 were included. The search strategy was designed to capture both preclinical and clinical studies.

### Summary of Included Studies

*The key findings and limitations of the included studies are summarized below:*

| Study | Design | Population | Key Findings | Limitations |
| --- | --- | --- | --- | --- |
| JAMA Network Open (2024) | Retrospective | Type 2 diabetes patients | No significant link between GLP-1RAs and thyroid cancer. | Limited to diabetes population. |
| OncLive (2023) | Preclinical | Endometrial cancer models | GLP-1 agonists may inhibit cancer cell proliferation. | Preclinical data; not yet tested in humans. |
| Healthline (2023) | Review | General population | Rodent studies show thyroid cancer risk, but human studies do not. | Lacks original data. |
| NCI (2024) | Clinical Trial | High-risk endometrial cancer patients | Testing semaglutide + progestin for cancer prevention. | Ongoing trial; results not yet available. |

### Inclusion and Exclusion Criteria

#### Studies were included if they

- Investigated the safety or efficacy of semaglutide in patients with a family history of thyroid or uterine cancer.
- Provided clear data on cancer risk or weight loss outcomes.

#### Studies were excluded if they

- Did not directly address the research question.
- Lacked sufficient methodological rigor (e.g., small sample size, no control group).

### Data Extraction and Analysis

Data from the included studies were extracted and summarized in the table above. Key outcomes included the incidence of thyroid or uterine cancer, weight loss efficacy, and adverse events. Due to the heterogeneity of the studies, a meta-analysis was not performed.

## SEMAGLUTIDE AND THYROID CANCER RISK

### Animal Studies

Available animal research shows that the use of semaglutide can increase the risk of developing thyroid C-cell tumours [1]. Based on these findings, the FDA inserted a boxed warning about thyroid C-cell tumors in semaglutide’s prescribing information.

### Human Studies

There is no evidence linking the use of semaglutide and the occurrence of thyroid cancer among other participants. A significant association between the use of GLP-1 receptor agonists and the development of thyroid cancers was not found in a retrospective study published by JAMA Network [3]. A Healthline article explained that rodent research indicated a higher risk, but human clinical trials did not find the same results [2]. However, some experts have raised concerns about the potential for overdiagnosis of thyroid cancer in patients using semaglutide, particularly in the context of increased screening and monitoring [7].

### Clinical Implications

Whenever medical attention is required, advanced-stage care is dreaded by all. In such circumstances, the people who have a case of extreme appetite ty single out a particular focus group, that is highly proactive in diagnosing medullary thyroid carcinoma or multiple neoplasm syndrome type two even though they can be linked to having little to no direct human evidence. Further studies have proposed that those partaking in the GLP-1 receptor enhancers tend to suffer from a higher diagnosis rate. This happens as there is concern that the worrying rate of cancer detection is inaccurate as the increased incidence might be the result of misdiagnosis rather than overly aggressive treatment escalation [6]. Additionally, some researchers suggest that the relationship between semaglutide and thyroid cancer risk may be more complex than previously thought, warranting further investigation into long-term safety [8].

## SEMAGLUTIDE AND UTERINE (ENDOMETRIAL) CANCER RISK

### Obesity and Endometrial Cancer

Excessive fat tissue increases the chances of women developing endometrial cancer. This is mainly true because women of reproductive age who are overweight have much more fat tissue, which leads to higher estrogen concentrations, the reason behind endometrial hyperplasia [4].

### Semaglutide’s Role

With the aid of semaglutide, the weight of these women can be brought under control, potentially reducing the number of women subjugated to endometrial cancer [5]. The link is hard to pin down clinically though.

### Preclinical and Clinical Evidence

Currently, the comparison between the combination of semaglutide with progestin versus progestin alone is being tested clinically for complying with endometrial cancer and making a cancer-free uterus and maintaining it [4]. In preclinical results, there is initial evidence that the development of cancer is the increase of disordered GLP-1 receptor agonists within estimable bounds collapsing the motility of cancer tissues in the endometrial model of cancer [5]. Semaglutide was found to inhibit cell proliferation in any sensible fashion much more than adjustable.

Notwithstanding these potential advantages, data on the long-term safety of semaglutide amongst patients with a family history of endometrial cancer remains absent. Considering the multifaceted interactions of hormones that contribute to endometrial cancer, it is crucial to conduct additional large-scale clinical studies [5].

## DISCUSSION

The present data does not point to any obvious relationship between semaglutide use and a higher risk of thyroid cancer development. Semaglutide is contraindicated in people with a personal or familial history of medullary thyroid carcinoma (MTC) or multiple endocrine neoplasia (MEN) syndrome because of some preclinical data and FDA warnings. Providers are encouraged to be judicious in assessing the patient’s history and risks before prescribing semaglutide for obesity management in these patients. Some research indicates that while semaglutide may not have a direct impact on thyroid cancer risk, more intensive surveillance of drug users could result in higher detection rates, which may be unrelated to cancer growth. [7]. Additionally, the long-term effects of semaglutide on thyroid are uncertain, and additional studies are necessary to establish its safety profile.

## Supporting information

Reporting checklist for systematic review (with or without a meta-analysis)

## Data Availability

All data produced in the present work are contained in the manuscript

https://jamanetwork.com/journals/jamanetworkopen/fullarticle/2820833

https://www.healthline.com/health/semaglutide-thyroid-cancer

https://www.onclive.com/view/preclinical-study-reveals-potential-role-for-glp-agonists-in-endometrial-cancer

https://www.healthline.com/health/semaglutide-thyroid-cancer

https://medlineplus.gov/druginfo/meds/a618008.html

https://www.onclive.com/view/preclinical-study-reveals-potential-role-for-glp-agonists-in-endometrial-cancer

https://www.health.com/thyroid-cancer-overdiagnosis-weight-loss-medications-ozempic-11680251

## Article Summary

### (Strengths and limitations of this study)

#### Strengths

1. Comprehensive Literature Search: This review utilized a systematic search strategy across multiple databases (PubMed, Google Scholar, and ClinicalTrials.gov) to ensure a broad and inclusive collection of relevant studies.
2. Focus on High-Risk Populations: The review specifically addresses the use of semaglutide in patients with a family history of thyroid or uterine cancer, a population that requires careful consideration due to potential risks.
3. Inclusion of Preclinical and Clinical Data: By including both preclinical and clinical studies, the review provides a balanced perspective on the potential risks and benefits of semaglutide in this context.
4. Clear Inclusion and Exclusion Criteria: The review employed strict criteria for study inclusion, ensuring that only relevant and methodologically sound studies were included in the analysis.
5. Up-to-Date Information: The review includes studies published up to 2024, providing the most current evidence available on the topic.

#### Limitations

1. Heterogeneity of Studies: The included studies varied in design, population, and outcomes, making it difficult to perform a meta-analysis or draw definitive conclusions.
2. Limited Human Data on Cancer Risk: While preclinical studies suggest a potential risk of thyroid C-cell tumors in rodents, human studies have not confirmed this risk. This discrepancy limits the ability to generalize findings to human populations.
3. Lack of Long-Term Data: Most studies included in this review were of short to medium duration, and long-term safety data on semaglutide use in patients with a family history of thyroid or uterine cancer are lacking.
4. Potential for Publication Bias: The review may be subject to publication bias, as negative or inconclusive results are less likely to be published and therefore may not have been included.
5. Focus on Specific Cancers: The review is limited to thyroid and uterine cancers, and the findings may not be applicable to other types of cancer or broader populations.

## Data Sharing Statement

*As required by BMJ Open, the following data sharing statement is provided*

### Data Availability

The data supporting the findings of this review are available from the corresponding author upon reasonable request.

### Data Sources

All data used in this review were extracted from publicly available studies and databases, including PubMed, Google Scholar, and ClinicalTrials.gov.

### Data Access

The full-text articles and clinical trial records referenced in this review can be accessed through their respective publishers or the ClinicalTrials.gov database.

### Data Sharing Restrictions

There are no additional restrictions on data availability beyond those imposed by the original studies or databases.

## CONCLUSION

Semaglutide is an effective treatment option for obesity management, although caution should be exercised in patients with a family history of thyroid or uterine malignancies. It is recommended that clinicians balance the expected weight loss benefits against the hypothetical dangers and supervise the patients very scrupulously.

## ETHICAL APPROVAL AND CONSENT

Ethical approval was not required for this review, as it is based on previously published studies and does not involve new human or animal subjects.

## AUTHOR CONTRIBUTIONS

The process of conceptualizing, gathering data, composing the original wording, conducting formal analysis with emphasis on epidemiology, and approving the manuscript as final steps is conducted by Mohammed Munshi.

Sabrina Jahangir is in charge of data collection, manuscript editing, and formatting.

*The design, analysis and interpretation of the data were equally contributed by both authors*.

## ACKNOWLEDGMENTS

The authors would like to thank Sumaiya Munshi and Abiyan Jahangir for their support and contributions to this review. We also acknowledge the researchers and institutions whose work was included in this systematic review.

